# The impact of COVID-19 restriction measures on loneliness among older adults in Austria

**DOI:** 10.1101/2020.09.08.20190397

**Authors:** Erwin Stolz, Hannes Mayerl, Wolfgang Freidl

## Abstract

**Background:** To halt the spread of COVID-19, Austria implemented a 7-week ‘shut-down’ of public life in March/April 2020 which was followed by a gradual withdrawal of these restriction measures in May/June 2020. We expect that the ensuing reduction in social contacts led to increased loneliness among older adults (60+).

**Methods:** We conducted three analyses to assess the association between COVID-19 public health restriction measures and loneliness: (1) A comparison between pre-pandemic (SHARE: 2013–2017) and pandemic (May 2020) levels of loneliness (UCLA-3 scale), (2) an analysis of the correlation between being affected by COVID-19 restriction measures and loneliness based on cross-sectional survey data from early May 2020, and (3) a longitudinal analysis of weekly changes in loneliness (Corona panel data) from late March to early June 2020.

**Results:** We found (1) loneliness levels to have increased in 2020 in comparison with previous years, (2) an association between the number of restriction measures older adults reported to be affected from and loneliness, and (3) that loneliness was higher during ‘shut-down’ compared to the subsequent re-opening phase, particularly among those who live alone.

**Discussion:** Our results provide evidence that COVID-19 restriction measures in Austria have indeed resulted in increased levels of loneliness among older adults. However, these effects seem to be short-lived, and thus we do not expect strong negative consequences for older adults’ mental health downstream. Nonetheless, effects of longer and/or repeated future restriction measures aiming at social distancing should be closely monitored.

## Introduction

The pandemic Coronavirus disease 2019 (COVID-19) represents an acute global health threat to human populations[1], with older adults particularly at risk for complications and death[2]. Lacking specific treatments or vaccines, exponential growth rates of COVID-19 cases, overburdened health care systems and high mortality rates in several hot spots early in the pandemic (e.g. Lombardy in Italy) have lead to unprecedented governmental public health measures to mitigate the spread of SARS-CoV-2. Many European countries restricted citizens movement and social contacts with stay-at-home orders, quarantines, and ‘shut-downs’ of public life over the course of several weeks or months[3]). In Austria, the ‘shut-down’ began on March 16th 2020 with the closing of stores, bars and restaurants, visiting bans in hospitals and nursing care homes, closing and/or controlling of borders, and regional quarantines. People were informed to stay at home and were allowed to leave the house only to go to work, buy groceries, help care-dependent others, or to take a walk, all while keeping at least one meter distance. In total, the strict ‘shutdown’ phase in Austria lasted for seven weeks, between mid March and the end of April. Since COVID-19 cases declined shortly after these measures were enacted – the maximum of around 9,000 confirmed cases was reached in early April – and COVID-19 deaths remained limited, most restrictions were again relaxed throughout May and early June 2020.

Although the enacted public health measures have successfully limited the spread of infection in Austria and lasted only for several weeks, there could be negative psychosocial side effects such as increased loneliness due to social isolation. This might affect older adults particularly, who are not only more vulnerable to COVID-19, but who, already before the pandemic, had an increased risk of loneliness due to widowhood, living alone, or mobility limitations[4, 5]. Particularly those who live alone might face an increased risk of loneliness, when social contacts with persons from outside the household are substantially reduced due to pandemic-related restriction measures[6, 7]. Loneliness can be defined as a subjective dissatisfaction with one’s social contacts or as a negative discrepancy between the desired and the perceived social relationships[8], and represents an important risk factor for poor physical and mental health outcomes[9, 10].

Whether levels of loneliness among older adults have increased substantively *and* sustainably during the pandemic, and whether this could result in negative physical and mental health outcomes downstream, is currently unclear. So far, only a few studies have assessed changes in loneliness among older adults with regard to the COVID-19 pandemic. Two studies[6, 11] from the U.S. – a country with high COVID-19 case numbers and heterogeneous policy responses – found that loneliness has increased during the pandemic compared to 2019 and early 2020. However, the effect size (Cohen’s d = 0.14) was quite small in the first study[6], and the sample small (n = 93) in the second study[11]. A study from the Netherlands[12] – a country which enacted a strict lockdown from mid March to the end of April 2020 and subsequent easing thereafter – reported substantive increases (Cohen’s d = 0.49) in loneliness between October/November 2019 and early May 2020 among more than 1,600 older adults (65+). Finally, a study among 1,070 older adults (aged 60–71) in Sweden[13] – a country which has remained open with few mandatory restrictions and more voluntary measures – showed no change in loneliness between the years 2015–2019 on the one hand, and (March-April) 2020 on the other hand. Varying results across countries are to be expected since the COVID-19 restriction measures also varied considerably across countries[3]. Thus evidence from more countries is needed in order to know what psychosocial side-effects can be expected if further strict restriction measures, even regional ones, are enacted in case of quickly rising COVID-19 cases throughout fall and winter 2020.

In this paper, we aim to assess the association between COVID-19 restriction measures and loneliness among older adults in Austria. Unfortunately, we are not aware of any well-timed individual-level survey data in Austria that would allow to compare the same older adults’ levels of loneliness before, during and after the COVID-19 ‘shut-down’. Thus, we resort to three indirect approaches: First, we use repeated cross-sectional data to compare population-level loneliness from previous years (2013–2017) from the Survey of Health, Ageing and Retirement in Europe (SHARE) with loneliness measured during the pandemic (May 2020) in Austria. Second, based on cross-sectional survey data from early May 2020, we assess whether older adults who reported to have been affected by more pandemic-related restrictions, also reported to be more lonely. Third, and finally, we analyse weekly changes in loneliness among a panel of older adults over the course of 10 weeks from late March to early June 2020, to see if these align with the course of the lifting of the COVID restriction measures.

## Methods

### Data

The first data set is a national, cross-sectional survey among community-dwelling older adults aged 60 years and above which was conducted at the behest of the authors by a professional survey agency, the Institute for Empirical Research (IFES, Vienna) during the first two weeks of May 2020. A majority of the interviews were conducted online (76%); telephone interviews were used particularly among older respondents (75+) to ensure adequate coverage. Respondents for the online interviews were randomly sampled from an offline and an online panel and invited via email to participate in the study; respondents for telephone interviews were randomly sampled using random-last digit screening. The response rate ranged between 40–45%, and a total of 557 interviews were conducted. Participants were informed about the study, the anonymity of all personal information and that they could end the interview at all times. All respondents provided consent to participate in the study. The study was approved by the Ethics Committee of the Medical University of Graz (EK-number 32–368 ex 19/20).

The second data set comes from the Austrian Corona Panel Project at the University of Vienna[14, 15]. Here, respondents were quota sampled from a pre-existing online access panel of another survey agency (Marketagent, Baden) based on key demographics (age, gender, region, municipality, size, and educational level). Online interviews were conducted weekly from late March (week 1 = 3/27/2020–3/30/2020) – i.e. two weeks after the lockdown in Austria began – to early June (week 10 = 5/29/2020–6/3/2020). The response rate for the first wave was 35.2%, which amounted to 1,541 interviews with respondents aged 14–75 years. 520 of these respondents were aged 60–75 years at the time of interview, but 25.4% respectively 24.2% did not provide valid answers to the two outcome measures at baseline, which reduced the effective number of respondents to 388 individuals with a total of 3,194 weekly observations. Respondents with non-valid answers at baseline were compared with respondents who provided a valid answer regarding socio-demographic characteristics.

Third, for a comparison of the extent of loneliness before and after the COVID-19 pandemic in Austria, we also rely on data from the Survey of Health, Ageing and Retirement in Europe (SHARE). Specifically, we use data from the fifth (March-November 2013)[16], sixth (Jannuary-November 2015)[17], and seventh (April-October 2017)[18] wave. SHARE is a harmonized cross-national survey study which provides information on community-dwelling older adults aged 50 years and over. For this study, we restricted the sample to Austrian participants aged 60 years and over at the time of the interview.

### Variables

In the cross-sectional data set, we used an established measure of loneliness, the three-item UCLA loneliness scale[19]. Specifically, respondents are asked how often they felt ‘a lack of companionship’, ‘left out’, or ‘isolated’ with answer categories ranging from ‘never’ (1) to ‘often’ (4). To keep the results comparable to the UCLA-3 loneliness scale used in SHARE, answer categories 1 (never) and 2 (rarely) were combined, so that there were only three answer categories. The resulting loneliness scale showed good internal consistency (Cronbach’s alpha = 0.78) and ranged from 3–9.

To measure the impact of COVID-19 restriction measures on people’s lives, we asked respondents in the cross-sectional survey whether they were (0 = no, 1 = yes) negatively affected from (1) restrictions regarding freedom of movement, (2) not being able to see children or grandchildren in person, (3) not being able to visit care-dependent older family members, (4) not being able to visit seriously or terminally ill family members, (5) not being able to participate in family celebrations/funerals, (6) restrictions regarding social (e.g. sport or cultural) activities, and (7) restrictions concerning visiting restaurants and cafes. These seven aspects were summed-up (range = 1–7, Cronbach’s alpha = 0.73) to quantify the exposure to COVID-19 restriction measures.

Further variables in the cross-sectional survey included sex (female/male), age (in years), completed high school education (no/yes), living alone (no/yes), depressive symptoms based on how strongly (1 = not at all, 5 = very strongly) respondents felt indifferent, depressed, worthless, hopeless, or suicidal in the last two weeks (Cronbach’s alpha = 0.85), and whether respondents had one or more chronic diseases (no/yes) potentially relevant for a COVID-19 prognosis (chronic respiratory disease, diabetes, cardiovascular disease and/or stroke, cancer, and diseases or therapies that suppress the immune system).

In the longitudinal Corona panel data, there are two single-item measures of loneliness: First, respondents were asked how strongly they agreed or disagreed with the following statement: ‘I miss contact with other people’, with answer categories ranging from ‘applies completely’ (1) to ‘does not apply at all’ (5). The variable was dichotomized, so that respondents who answered with ‘applies completely’, ‘rather applies’ or ‘partly applies’ were coded as having missed contact (1), and those who answered that this does ‘not’ or ‘rather not’ apply were coded as 0. Second, participants were asked how often they felt lonely during the last week. Possible answer categories included ‘never’ (1), ‘on some days’ (2), ‘multiple times a week’ (3), ‘almost everyday’ (4), and ‘everyday’ (5). We categorized respondents who answered ‘never’ as not having felt lonely (= 0) during last week, and all others as having felt lonely (= 1).

Control variables in the Corona panel data included the same socio-demographics as in the cross-sectional dataset, i.e. sex (female/male), age (in years), completed high school education (no/yes), and living alone (no/yes), and whether respondents had one or more chronic diseases (cardiovascular disease, diabetes, hepatitis B, chronic obstructive lung disease, chronic kidney disease, or cancer) relevant for a COVID-19 prognosis.

### Statistical analysis

We first describe the demographic characteristics of the cross-sectional and the longitudinal sample. Second, we compare population-level loneliness (UCLA-3) between three pre-pandemic waves of SHARE Austria and our within-pandemic cross-sectional survey study. Third, we used linear regression models to assess the association between how strongly respondents were affected by COVID-19 restriction measures and their level of loneliness based on the cross-sectional sample. Analyses were adjusted for all control variables, and we assessed whether the effect of restriction measures was moderated by living alone. Fourth, we used mixed logistic regression models with random intercept and random slope (week) effects to assess how loneliness changed over the course of 10 weeks in the longitudinal data set. Since loneliness might change non-linearly over time as the lockdown phase was followed by subsequent re-opening phase, we used a flexible thin-plate regression spline[p.215 20] to estimate the effect of time. To assess whether change in loneliness was moderated by living alone, we tested for interaction effects between living alone and (non-linear) time. Model comparisons were based on the Watanabe Akaike Information Criterion (WAIC). All models were estimated with ‘brms’ (v.2.11.1)[21], a front-end for ‘RStan’ (v.2.19.2)[22] in R: A language and environment for statistical computing (v.3.6.3)[23].

### Data and code availability

Data from the Survey of Health, Ageing and Retirement in Europe (SHARE) is freely accessible for researchers upon registration
^1^
. The cross-sectional survey data gathered by IFES at the behest of the authors is available upon request, and will be made generally available for scientific use via the Austrian Social Science Data Archive (AUSSDA) later in 2020. The longitudinal data from the Corona panel is already available via AUSSDA
^2^
. The R-Markdown code reproducing all analyses and results is also available online
^3^
.

## Results

Sociodemographic characteristics were comparable in the cross-sectional and the longitudinal sample (eTable 1 in the appendix). In the cross-sectional survey from early May 2020, respondents were asked whether or not they are currently negatively affected by seven aspects of COVID-19 restriction measures aiming to reduce social contact. On average, respondents were negatively affected by 3.4 aspects (SD = 1.6) of pandemic-related restrictions. More specifically, 80.7% stated they were affected negatively by not being able to visit restaurants and bars, 72.4% by not being able to participate in social, sport or cultural activities, 62.6% by restricted freedom of movement, 57.8% by not being able to see children or grandchildren in person, 36.0% by not being able to participate in family celebrations (including funerals), 18.5% by not being able to visit care-dependent older adults, and 9.4% by not being able to visit seriously or terminally ill family members.

The median loneliness value in the UCLA-scale was 4 based on our cross-sectional sample from May 2020. In comparison, the median value of the UCLA scale in SHARE Austria from 2013, 2015, and 2017 was 3, i.e. lower by one point (on scale from 3–9). This also shows (Panel A in Figure 1) in the distribution of values in the UCLA-scale (range = 3–9) over time. The repeated cross-sectional data from SHARE shows little change over several years (2013–2017), but there is a substantial increase between these years and 2020. This increase in loneliness is also visible at the item-level: For example, in 2017, 93% of the respondents in SHARE answered ‘hardly ever or never’ to the question ‘how much of the time do you feel isolated from others?’, 6% answered ‘some of the time’ and only 1% answered with ‘often’. In contrast, in May 2020, 64% answered that they ‘hardly ever’ or ‘never’ felt isolated, but 27% answered ‘some of the time’ and 9% ‘often’.

The bivariate Pearson correlation coefficient between the number of pandemic-related restrictions and loneliness in the cross-sectional dataset was 0.34 (n = 551, p< 0.001). Results from the linear regression model (eTable 2) confirm the association between the number of pandemic-related restrictions respondents were affected by and their level of loneliness (Panel B of Figure 1). Despite adjustment for a number of potential confounders, including living alone, the presence of chronic diseases, and depressive symptoms, we found that respondents who reported to be affected by more pandemic-related restrictions, also felt more lonely (unadjusted effect size = 0.36 (CI-95 = 0.28–0.45); adjusted effect size = 0.28 (CI-95 = 0.20–0.35). The moderation effect of living alone did not improve model fit (without interaction effect: WAIC = 1,871 (SE = 55); with interaction effect: WAIC = 1,873 (SE = 55); WAIC-weight = 76% for the model without interaction effect).

Missing information for the two outcomes at baseline in the Corona panel data was related to sex and educational level, but not age, living alone or having chronic diseases. Specifically, we found that women (felt lonely: women = 31.5%, men = 18.3%; *χ*^2^ = 12.05, df = 1, p< 0.001; missed social contact: women = 29.7%, men = 17.8%; *χ*^2^ = 9.98, df = 1, p = 0.001) and respondents who completed high school level education (felt lonely: high-school education = 40.0%, no high-school education = 21.6%; *χ*^2^ = 15.5, df = 1, p< 0.001; missed social contact: high-school education = 39.1%, no high-school education = 20.3%, *χ*^2^ = 16.54, df = 1, p< 0.001) were more likely to *not* provide a valid answer to the two items on loneliness compared to men and those with a lower level of education.

The trend in the reported proportion of the two loneliness items from the Corona panel during the lock-down phase and the subsequent re-opening phase are shown in Panel C (Figure 1). We find that missed social contacts showed a substantive reduction after the ‘shut-down’ ended (mean value during ‘shut-down’ = 56.6%, mean value during re-opening phase = 45.8%), while ‘feeling lonely’ decreased less (mean value during ‘shut-down’ = 32.4%, mean value during re-opening phase = 28.6%) over the same period. Results from the logistic mixed regression model on how loneliness changed over time during the pandemic are shown in Panels D and E of Figure 1, and eTable 3. The change in the predicted probability of missing social contacts confirms the strong decrease shortly after the restrictions were lifted. The moderation effect of living alone did not improve model fit (WAIC = 2,617 (SE = 86) vs. WAIC = 2,619 (SE = 85), WAIC weight = 0.71 for the model without interaction effect). Living alone, however, was relevant for the trend in feeling lonely (WAIC = 2,105 (SE = 86) vs. WAIC = 2,113 (SE = 86); WAIC weight = 97% for the model with moderation effect): While there is little change among older adults who lived with others over time, the probability of feeling lonely decreased substantively (from 30–40% during lockdown to 10% at the end of the observation period) among older adults who lived alone.

**Figure 1.**
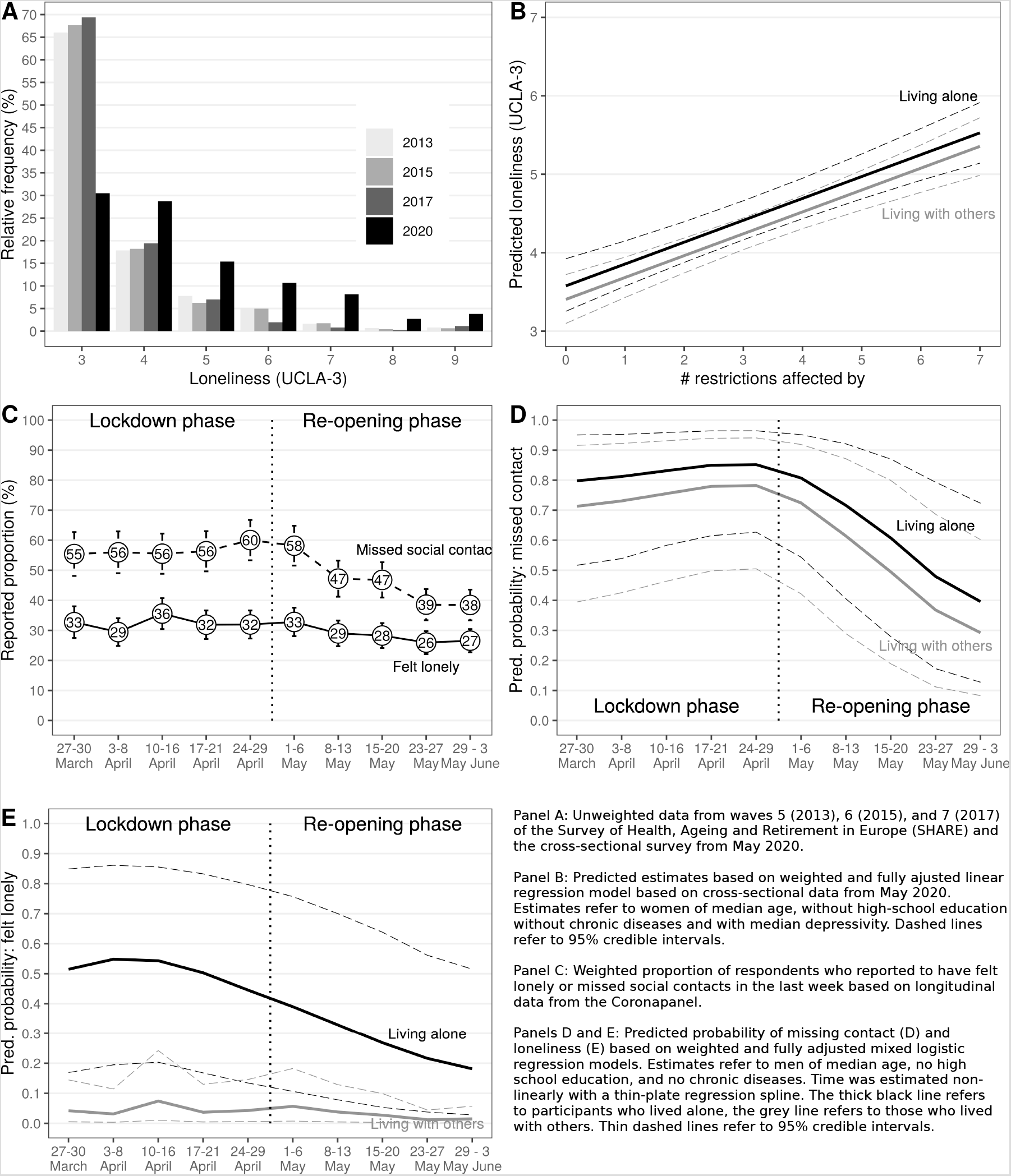
Results from analyses based on cross-sectional (A,B) and longitudinal (C-E) data on loneliness.

## Discussion

In this analysis, we assessed the association between COVID-19 restriction measures and loneliness among older adults in Austria. In summary, we found (1) loneliness levels to have increased in 2020 in comparison with earlier years, (2) an association between the number restriction measures older adults were affected by and loneliness, and (3) that loneliness was higher during lockdown compared to the subsequent re-opening phase, particularly among those who live alone. Together, these results suggest that COVID-19 restriction measures in Austria have resulted in increased levels of loneliness among older adults.

Our results are compatible with findings of studies from the U.S.[6, 11] and one study from the Netherlands[12], which also found increased levels of loneliness during the COVID-19 pandemic. The plausible impact of COVID-19 restriction measures on loneliness among older adults is further corroborated by findings from Sweden[13], where restriction measures were limited and loneliness levels did not change. Different to previous work, we measured the number of COVID-19 restriction measures older adults were affected by also directly. We found that a substantial association between the number of COVID-19 restriction measures participants were affected by and loneliness under adjustement for a number of control variables, which, importantly, included depressive symptoms. Also different to previous studies, we used intensive (i.e. weekly) longitudinal data to follow how reported loneliness responded to the lifting of COVID-19 restriction measures. We found a close temporal correspondence: Soon after the restriction were relaxed, older adults were considerably less likely to report missing social contacts, and, for those who live alone, also to report feeling lonely. Therefore, we expect this to be a generally rather short-lived effect, so that a few weeks after restrictions are relaxed or lifted, the prevalence of loneliness should return to the levels before strict restriction measures were enacted. Although we therefore do not expect strong negative mental health consequences downstream due to restriction-related loneliness for most older adults[12], the increased distress of even short-term loneliness might be enough to clinically worsen mental health among particularly vulnerable older adults. More research is needed to address and mitigate negative mental health consequences of COVID-19 restriction measures among particularly vulnerable older adults, for example those with pre-existing mental health issues[24, 25]. Also, although we consider the effect of the restriction measures on loneliness to be short-lived, this does not necessarily mean that loneliness levels will go back to the pre-pandemic long-term average level. Fear of infection, caution, and voluntary and preventive limiting of social contacts may operate above and beyond public health restriction measures and may result in negative mental and physical health consequences for older adults who perceive themselves – or a are perceived by others – as particularly vulnerable to COVID-19. Finally, it is unclear whether the suggested lagged dose-effect relationship between COVID-19 restriction measures and levels of loneliness is not, at least partly, due to the novelty of the situation. The effects on loneliness, and subsequent mental health issues, might be both more long-lasting and severe if future restriction measures, even local ones, are enacted over longer time periods and/or repeatedly, in other words if social distancing and/or isolation becomes the ‘new normal’ over a potentially multi-year-long period until a vaccine or treatment for COVID-19 become available. There are several noteworthy limitations to our analysis. First, to the best of our knowledge, there is no population-representative, individual-level longitudinal data available that covers loneliness among the same older adults before the pandemic, during the lockdown phase and after the lockdown phase in Austria. In consequence, we had to resort to three more indirect analyses: (1) a comparison of cross-sectional survey data during the lockdown with pre-pandemic repeated cross-sectional data from SHARE, (2) an assessment of the association between being affected by COVID-19 restriction measures and loneliness, and (3) an analysis of weekly changes in loneliness during and after the lockdown phase in Austria. Although these analyses cannot substitute for consistent, high-quality, individual-level longitudinal survey data, we consider them the best available approximations to assess the likely impact of COVID-19 restriction measures on loneliness among older adults in Austria. Second, in contrast to the cross-sectional data analysis where an established instrument for loneliness was used, in the longitudinal data, loneliness was measured only with two single ordinal items, which were further dichotomized during the analysis. Thus, we could not differentiate between different degrees of loneliness in the longitudinal data analysis. Third, it is unclear whether both the cross-sectional and longitudinal sample are representative for the population of older adults regarding loneliness. First, older adults with severe health impairments – which are more likely to feel lonely due to loss of partners, siblings and other peers – are less likely to participate in survey studies[26], and residents of care-homes, which were particularly affected by visiting bans[27], were not included in the sampling strategy at all. In the Corona panel specifically, about one quarter of the respondents did not provide valid information on loneliness, and this was more prevalent among women and those with higher educational qualification, which may bias our results. Also, in the Corona panel, there were no participants aged 76 year or older as these are difficult to reach with online interviews. However, in the cross-sectional survey which combined online- and telephone-interviews and included older adults up to age 89, we found no difference due to the interview mode (telephone vs. online interview) in loneliness.

In conclusion, our results provide evidence that COVID-19 restriction measures in Austria indeed increased levels of loneliness among older adults. However, these effects are likely short-lived, and thus we do not expect strong negative consequences for older adults’ mental health. Nonetheless, effects of longer and/or repeated future restriction measures aiming at social distancing should be closely monitored.

## Data Availability

Data from the Survey of Health, Ageing and Retirement in Europe (SHARE) is freely accessible for researchers upon registration. The cross-sectional survey data gathered by IFES at the behest of the authors is available upon request, and will be made generally available for scientific use via the Austrian Social Science Data Archive (AUSSDA) later in 2020. The longitudinal data from the Corona panel is already available via AUSSDA. The R-Markdown code reproducing all analyses and results is also available online.

http://www.share-project.org/data-access.html

https://data.aussda.at/dataset.xhtml?persistentId=doi:10.11587/28KQNS

https://osf.io/ck76a/

## Footnotes

1 http://www.share-project.org/data-access.html

2 https://data.aussda.at/dataset.xhtml?persistentId=doi:10.11587/28KQNS

3 https://osf.io/ck76a/

## Appendix

**eTable 1:** Sample characteristics

|  | Cross-sectional sample | Longitudinal sample (baseline) |
| --- | --- | --- |
| Female sex: % (n) | 54.3 (292) | 54.3 (279) |
| Age: median, IQR (n) | 70 (10, 557) | 67 (9, 520) |
| High school education: % (n) | 20.0 (244) | 20.0 (110) |
| Living alone: % (n) | 29.5 (170) | 30.4 (165) |
| Chronic disease: % (n) | 40.0 (225) | 34.4 (179) |

Descriptives based on weighted data, (n) = sample size, IQR = interquartile range covering data from the 25th to the 75th quantile

**eTable 2:**
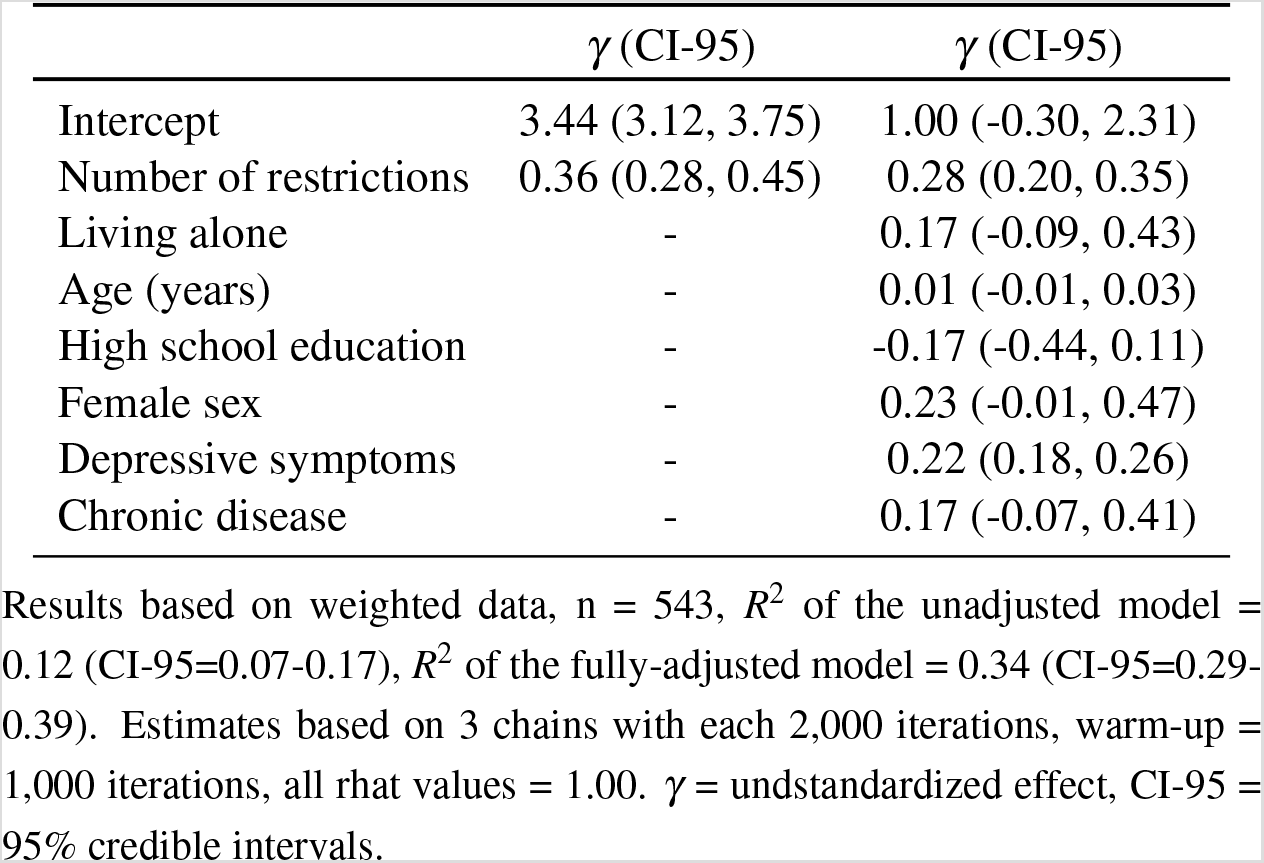
Results from linear regression model based on cross-sectional data

| | $\gamma$ (CI-95) | $\gamma$ (CI-95) |
| --- | --- | --- |
| Intercept | 3.44 (3.12, 3.75) | 1.00 (-0.30, 2.31) |
| Number of restrictions | 0.36 (0.28, 0.45) | 0.28 (0.20, 0.35) |
| Living alone | - | 0.17 (-0.09, 0.43) |
| Age (years) | - | 0.01 (-0.01, 0.03) |
| High school education | - | -0.17 (-0.44, 0.11) |
| Female sex | - | 0.23 (-0.01, 0.47) |
| Depressive symptoms | - | 0.22 (0.18, 0.26) |
| Chronic disease | - | 0.17 (-0.07, 0.41) |

Results based on weighted data, $n = 543$ , $R^2$ of the unadjusted model = 0.12 (CI-95=0.07-0.17), $R^2$ of the fully-adjusted model = 0.34 (CI-95=0.29-0.39). Estimates based on 3 chains with each 2,000 iterations, warm-up = 1,000 iterations, all rhat values = 1.00. $\gamma$ = undstandardized effect, CI-95 = 95% credible intervals.

**eTable 3:** Results from mixed logistic regression model based on longitudinal data

| | Missed social contact<br>$\gamma$ (CI-95) | Felt lonely<br>$\gamma$ (CI-95) |
| --- | --- | --- |
| POPULATION-LEVEL EFFECTS |  |  |
| Intercept | -3.85 (-9.65, 1.88) | -1.60 (-8.26, 4.93) |
| Living alone | 0.51 (-0.34, 1.40) | 3.20 (2.24, 4.24) |
| Female sex | 1.16 (0.34, 1.97) | 0.73 (-0.26, 1.69) |
| Age (years) | 0.05 (-0.03, 0.13) | -0.04 (-0.14, 0.05) |
| High school education | -0.32 (-1.35, 0.74) | 0.10 (-1.13, 1.31) |
| Chronic disease | 0.60 (-0.24, 1.43) | 1.52 (0.56, 2.50) |
| Week | -4.27 (-15.03, 8.24) | - |
| Week*living with others | - | -5.64 (-12.87, 17.33) |
| Week*living alone | - | -1.00 (-12.87, 17.33) |
| INDIVIDUAL-LEVEL EFFECTS |  |  |
| Intercept | 4.08 (3.38, 4.91) | 3.82 (3.06, 4.69) |
| Week | 0.38 (0.31, 0.45) | 0.35 (0.26, 0.45) |
| Corr. Intercept*week | -0.54 (-0.72, -0.31) | -0.19 (-0.50, 0.14) |
| SMOOTH TERMS |  |  |
| Week | 4.81 (1.93, 10.87) | - |
| Week*living with others | - | 21.29 (3.66, 43.38) |
| Week*living alone | - | 3.70 (0.35, 10.29) |

Results based on weighted data, n-individuals = 378, n-observations = 3,117, $R^2$ of the fully-adjusted model of missed social contact was 0.18, for felt lonely it was 0.22. Smooth terms were estimated using a thin-plate regression spline and provide information about the wiggleness. Estimates based on 3 chains with each 2,000 iterations, warm-up = 1,000 iterations, all rhat values = 1.00. $\gamma$ = unstandardized effect on logit scale, CI-95 = 95% credible intervals.

